# Genome-wide association analysis reveals extensive genetic overlap between mood instability and psychiatric disorders but divergent patterns of genetic effects

**DOI:** 10.1101/2021.07.16.21260608

**Authors:** Guy Hindley, Kevin S. O’Connell, Zillur Rahman, Oleksandr Frei, Shahram Bahrami, Alexey Shadrin, Margrethe Collier Høegh, Weiqiu Cheng, Naz Karadag, Aihua Lin, Linn Rødevand, Chun C. Fan, Srdjan Djurovic, Trine Vik Lagerberg, Anders M. Dale, Olav B. Smeland, Ole A. Andreassen

## Abstract

Mood instability (MOOD) is a transdiagnostic phenomenon with a prominent neurobiological basis. Recent genome-wide association studies found significant positive genetic correlation between MOOD and major depression (DEP) and weak correlations with other psychiatric disorders. We investigated the polygenic overlap between MOOD and psychiatric disorders beyond genetic correlation to better characterize putative shared genetic determinants. Summary statistics for schizophrenia (SCZ, n=105,318), bipolar disorder (BIP, n=413,466), DEP (n=450,619), attention-deficit hyperactivity disorder (ADHD, n=53,293) and MOOD (n=363,705), were analysed using the bivariate causal mixture model and conjunctional false discovery rate methods to estimate the proportion of shared variants influencing MOOD and each disorder, and identify jointly associated genomic loci. MOOD correlated positively with all psychiatric disorders, but with wide variation in strength (rg=0.10-0.62). Of 10.4K genomic variants influencing MOOD, 4K-9.4K were estimated to influence psychiatric disorders. MOOD was jointly associated with DEP at 163 loci, SCZ at 110, BIP at 60 and ADHD at 25, with consistent genetic effects in independent samples. Fifty-three jointly associated loci were overlapping across two or more disorders (transdiagnostic), seven of which had discordant effect directions on psychiatric disorders. Genes mapped to loci associated with MOOD and all four disorders were enriched in a single gene-set, “synapse organization”. The extensive polygenic overlap indicates shared molecular underpinnings across MOOD and psychiatric disorders. However, distinct patterns of genetic correlation and effect directions of shared loci suggest divergent effects on corresponding neurobiological mechanisms which may relate to differences in the core clinical features of each disorder.

## INTRODUCTION

Mood instability (MOOD) is a psychological construct defined as a tendency to experience frequent, rapid fluctuations of intense affect and an inability to regulate these fluctuations or their behavioural sequelae (1). The concept was first described in people with borderline personality disorder and is a central component of the disorder (2). While present in approximately 14% of the general population (3), it is also overrepresented in several other psychiatric disorders, including schizophrenia (SCZ), bipolar disorder (BIP), depression (DEP), and attention-deficit hyperactivity disorder (ADHD) (4–7) Furthermore, MOOD is a predictor and trait-marker for both DEP and BIP (6,8–10), and is associated with suicidality and poor treatment outcomes in multiple disorders (4,11).

There is mounting evidence supporting a prominent neurobiological basis to MOOD. Firstly, twin studies have estimated 25% and 40% heritability for affect intensity and affective lability respectively, central components of MOOD (12). Secondly, symptoms mirroring MOOD can be caused by seizure activity or localised brain lesions, typically involving the pre-frontal cortex, the temporal lobe and the diencephalon (13). Thirdly, neuroimaging, behavioural, cognitive and electrophysiological studies have reported an array of neurobiological correlates, of which alterations in amygdala activation and connectivity between the ventromedial prefrontal cortex, amygdala and anterior cingulate cortex are the most convincing (14). In combination with its clinical significance, MOOD therefore represents a promising transdiagnostic therapeutic target that could be leveraged to develop novel treatments and inform personalized psychiatric treatment, consistent with the Research Domain Criteria framework (15). Despite this, questions remain over MOOD’s neurobiological and phenomenological consistency across and within diagnostic groups, particularly in disorders such as SCZ, which is classically associated with reduced affective expression despite increased MOOD (2,16).

An improved understanding of the shared genetic basis of MOOD and different psychiatric disorders may provide insights into these questions. Two large-scale GWASs of MOOD in the UK Biobank have previously identified 46 genomic loci and strong positive genetic correlations with depression, but weak positive correlations with SCZ, BIP and ADHD (17,18). This has implicated several genes in MOOD which are also implicated in psychiatric disorders, including *PLCL1* in SCZ, *PLCL2* in BIP and *NEGR1* in DEP (18–21). Nonetheless, much of the genetic basis for MOOD and psychiatric disorders remains unexplained and individual loci linked to both have yet to be examined systematically (18,22). Furthermore, the identification of overlapping loci might help to disentangle the effects of different risk loci on the diverse phenomenology of psychiatric disorders and highlight neurobiological pathways with therapeutic potential (15).

To this end, we applied state-of-the-art statistical tools to summary statistics from GWAS of MOOD, SCZ, BIP, DEP and ADHD (18–21,23). We first used the bivariate causal mixture model (MiXeR) to estimate the total number of trait-influencing variants shared between MOOD and psychiatric disorders (24). Since MiXeR quantifies total genetic overlap and is unable to identify shared genomic loci, we next employed the conjunctional false discovery rate (conjFDR) method to discover loci jointly associated with MOOD and each psychiatric disorder beyond genome-wide significance (25). Unlike genetic correlation, which provides an aggregate measure for the balance of variants with concordant and discordant effects on two phenotypes, MiXeR and conjFDR are able to identify genetic overlap regardless of effect direction (26). These methods complement genetic correlation to provide a more comprehensive overview of the genetic relationships between phenotypes. Given MOOD’s increased prevalence across multiple diagnostic categories, we also aimed to identify loci that were common to MOOD and more than one psychiatric disorder, representing “transdiagnostic” MOOD loci. Finally, the conjFDR method also leverages cross-phenotype enrichment to boost the power to identify novel genomic loci for each phenotype, thus contributing to efforts to unravel the “missing” heritability of psychiatric disorders (22,25).

## METHODS AND MATERIALS

### Samples

We acquired summary statistics from a GWAS of MOOD in the UK Biobank (n=363,705) (18). MOOD was assessed by a yes/no questionnaire item “does your mood often go up and down?” (18). While this only captures “frequent fluctuations of affect” and no other features of MOOD, e.g. affect intensity (1), a positive response has been found to be 2.5 and 14.3 times more prevalent in people with DEP and BIP compared to controls, demonstrating its clinical relevance (8). The SCZ sample comprised a meta-analysis of CLOZUK and Psychiatric Genomics Consortium (PGC) consisting of 40,675 cases and 64,643 controls (19). The DEP sample was a meta-analysis of PGC and 23andMe, Inc. samples comprising a total of 121,198 cases and 329,421 controls (21). BIP and ADHD samples were acquired from the latest PGC GWAS, comprising 41,917 cases and 371,549 controls for BIP (27) and 19,099 cases and 34,194 controls for ADHD (23). All samples were of European descent. We also included height as a non-psychiatric comparator (n=709,706) (28). The PGC East Asian SCZ sample (cases = 22,778, controls = 35,362) (29) and FinnGenn BIP (cases = 4,501, controls = 192,220) and DEP (cases = 17,794, controls = 156,611) samples (30) were used for replication (*Supplementary methods*). The Norwegian Institutional Review Board: Regional Committees for Medical and Health Research Ethics (REC) South-East Norway evaluated the current protocol and found that no additional ethical approval was required because no individual data were used.

### Data analysis

MiXeR v1.3 was applied to MOOD and each of SCZ, BIP, DEP, ADHD, and height (24). MiXeR first uses a univariate gaussian mixture model to quantify the polygenicity of each trait from GWAS summary statistics, expressed as the number of ‘trait-influencing’ variants (also referred to as ‘causal’ variants). Next, a bivariate gaussian mixture model is constructed to quantify the additive genetic effect of four components: 1) variants not influencing either phenotype; variants uniquely influencing either the 2) first or 3) second phenotype and 4) variants influencing both phenotypes. Results are visualised as a Venn diagram. MiXeR also calculates the genetic correlation between phenotypes and predicts the proportion of shared variants with concordant effect direction on both phenotypes. Estimates and standard errors are calculated by performing 20 iterations using 2 million randomly selected SNPs for each iteration, followed by random pruning at an linkage disequilibrium threshold of r^2^=0.8. Model fit is based on likelihood maximization of signed test statistics (GWAS z-scores), evaluated by the Akaike Information Criterion (AIC) and demonstrated with modelled versus observed conditional quantile-quantile (Q-Q) plots.

We next employed conjFDR to identify SNPs jointly associated with MOOD and each psychiatric disorder, which has been described previously in detail (25,31). Briefly, conditional Q-Q plots were constructed to visualise cross-trait polygenic enrichment of SNP associations between MOOD and each psychiatric disorder (*Supplementary methods*). Cross-trait enrichment was leveraged within a Bayesian statistical framework to boost the power to discover shared genetic loci beyond genome-wide significance. Computed as the maximum of two mutual conditional FDR values (*Supplementary methods*), the conjFDR value provides an estimate for the posterior probability that a SNP is not associated with either trait or both traits. SNPs with a conjFDR<0.05 were assigned statistical significance.

The consistency of genetic effects in independent samples was evaluated using an *en-masse* sign concordance test (*Supplementary methods*) (32–34).

### Genomic loci definition

Independent genomic loci jointly associated with MOOD and each psychiatric disorder were defined using the FUMA protocol (35). Significant, independent SNPs were defined as conjFDR<0.05 and r^2^<0.6. Lead SNPs were chosen if they were in approximate linkage equilibrium with each other (r^2^<0.1). Transdiagnostic loci were defined as physically overlapping loci which shared at least one candidate SNP with conjFDR≤0.05 across two or more MOOD/psychiatric disorder conjunctional analyses. Effect directions within transdiagnostic loci were evaluated by comparing effect sizes of the SNP with the lowest maximum conjFDR value within the overlapping region from each MOOD/psychiatric disorder analysis, defined as the “transdiagnostic lead SNP”. LD data was calculated using the European population of the 1000 genomes project reference panel (36).

### Functional annotation

Candidate SNPs, defined as any SNP within each jointly associated genomic locus with a conjFDR value <0.10 and an LD r2≥0.6 with an independent significant SNP, were functionally annotated using FUMA (35). A lower conjFDR threshold for candidate SNPs was employed to maximise the probability that putative causal SNPs are captured for functional annotation, consistent with previous primary GWAS (33) and conjFDR studies (37). SNPs were mapped to putative causal genes using three strategies: a) positional mapping b) expression quantitative trait locus (eQTL) mapping and c) chromatin interaction mapping(35). We defined a subset of “credible mapped genes” as those that were mapped by all three strategies. We conducted Gene Ontology gene-set analyses using FUMA (35) on credible mapped genes for each MOOD/psychiatric disorder pair. See *Supplementary methods* for further details.

### Data availability

All GWAS summary statistics are publicly available besides 23andMe DEP data (*Supplementary methods*). The code for all analyses can be accessed at https://github.com/precimed.

## RESULTS

### Using MiXeR to estimate total polygenic overlap

Univariate MiXeR demonstrated MOOD to be highly polygenic, with 10.4K (SD=0.4K) variants predicted to influence MOOD, comparable to the complex polygenic architectures of psychiatric disorders (*Supplementary table 1, Supplementary figure 1*).

Bivariate MiXeR analysis revealed substantial overlap between MOOD and all four disorders (*Figure 1a, Supplementary table 1*), both in the presence of moderate positive genetic correlation (DEP and ADHD) and minimal genetic correlation (SCZ and BIP). This occurs due to a pattern of mixed effect directions among shared variants, i.e. a balance of variants with concordant and discordant effects on each trait cancel each other out resulting in minimal correlation despite extensive polygenic overlap. For example, the overlap between SCZ and MOOD was particularly striking, with 9.4K (SD=0.4K) shared variants, representing 97% variants influencing SCZ and 90% variants influencing MOOD, despite weak positive genetic correlation (r_g_=0.11, SE=0.0089). There was also weak positive genetic correlation (r_g_=0.10, SE=0.0096) but fewer shared variants between BIP and MOOD, with 7.8K (SD=0.6K) shared variants which represented smaller proportions of trait-influencing variants (91% and 75% for BIP and MOOD respectively). The proportions of shared variants predicted to have concordant effects on MOOD and each of SCZ (54%, SD=0.4%) and BIP (57%, SD=0.5%) were consistent with extensive overlap and weak genetic correlation.

**Figure 1:**
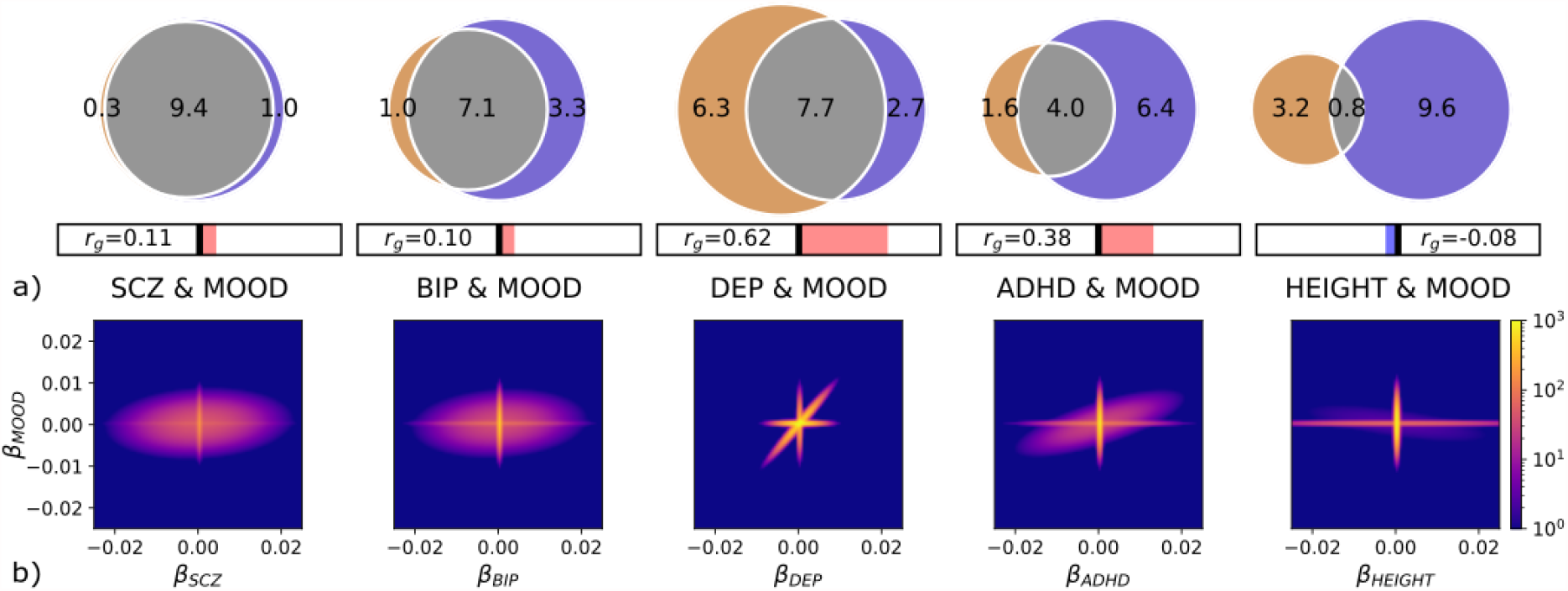
***a)*** Total number of shared variants between mood instability (MOOD, blue) and schizophrenia (SCZ), bipolar disorder (BIP), major depression (DEP), attention-deficit hyperactivity disorder (ADHD) and height as estimated by MiXeR. Venn diagrams representing the proportion of unique and shared variants associated with MOOD and each of SCZ, DEP, BIP, ADHD and height. Polygenic overlap is represented in grey. The numbers indicate the estimated quantity of variants in thousands per component that explains 90% of SNP heritability for each phenotype. The size of the circle reflects the extent of polygenicity for each trait. Genetic correlation (rg) is represented in the horizontal bars beneath the Venn diagrams. Right of the central bar (red) indicates positive rg and left of the central bar (blue) indicates negative rg. ***b)*** MiXeR density plots illustrating the effect sizes of variants (β, x and y axes) influencing MOOD and each of SCZ, BIP, DEP, ADHD, and height respectively, with respect to the number of variants (colour scale, blue to yellow). Extensive polygenic overlap of concordant and discordant variants is observed for MOOD and SCZ and BIP. The plots of MOOD and ADHD and DEP also illustrate extensive polygenic overlap, but most variants have concordant effects. The plot of MOOD and height indicates that most variants influencing each trait have little to no effect on the other.

In comparison, DEP (r_g_=0.62, SE=0.011) and ADHD (r_g_=0.38, SE=0.012) possessed stronger positive genetic correlations with MOOD (replicating previous findings in DEP) (18). A total of 7.7K (SD=0.3K) variants were estimated to be shared between DEP and MOOD, representing 55% DEP-influencing variants and 74% MOOD-influencing variants. The high number of DEP-specific variants relative to the other disorders (6.3K, SD=0.5K, 45%) was likely due to DEP’s extensive polygenicity (14K, SD=0.6K) and may relate to the clinical heterogeneity of the disorder (24,38). ADHD was found to share 4K (SD=0.6K) variants, representing 71% ADHD influencing variants and 38% MOOD influencing variants. The high proportion of MOOD-specific variants (6.4K, 0.6K, 62%) is related to ADHD’s lower polygenicity (5.6K, SD=0.4K) relative to MOOD. Consistent with the stronger positive genetic correlations, there were higher proportions of shared variants predicted to have concordant effects, with 94% (SD=2.8%) concordant for DEP and MOOD and 77% (SD=6%) for ADHD and MOOD. Given the extensive polygenic overlap across phenotypes, we applied the MiXeR model to height and MOOD as a non-psychiatric comparator. MiXeR estimated 0.8K (0.2K) shared variants and minimal negative correlation (r_g_=-0.08, SE=0.0083). AIC and conditional QQ plots to assess MiXeR model fit are described in *Supplementary results*.

The relationship between the number of shared variants and genetic correlation is illustrated in density plots in *Figure 1b*, in which the effect of each variant on MOOD is plotted against its effect on each psychiatric disorder and height. For SCZ and BIP, a large proportion of variants effect both phenotypes (oval) but these are distributed evenly between regions indicating concordant effects (top-right and bottom-left quadrants) and discordant effects (top-left and bottom-right quadrants). The effects of overlapping SNPs cancel each other out leading to weak genetic correlation despite substantial overlap. For DEP and ADHD, most variants have concordant directions, illustrated by the preponderance of variants in the top-right and bottom-left quadrants. This results in polygenic overlap and stronger positive genetic correlations. The MOOD and height subplot reveals that most variants affecting one trait do not influence the other and vice versa. Almost all associated variants are therefore plotted close to β=0 for one or the other phenotype (horizontal and vertical lines), indicating minimal genetic overlap and weak genetic correlation.

### Identifying genomic loci shared between MOOD and psychiatric disorders

We computed conjFDR values for each SNP present in both primary GWASs. The conjFDR value is interpreted as a conservative estimate for the probability that a given SNP is not associated with either phenotype. By leveraging cross-trait enrichment and employing a Bayesian statistical framework (*Supplementary figures 3-4*), conjFDR identifies jointly associated loci beyond genome-wide significance.

At conjFDR<0.05, MOOD was jointly associated with SCZ at 102 independent genomic loci, BIP at 60 loci, DEP at 163 loci, and ADHD at 28 loci (*Table 1, Supplementary table 2*). Among these, 246 were novel in MOOD, 26 in SCZ, 22 in BIP, 92 in DEP and 12 in ADHD, demonstrating conjFDR’s ability to boost the power to discover novel loci. On comparing the effect direction of jointly associated lead SNPs, 58.9% (60/102) were concordant for SCZ and MOOD, 65.0%% (39/60) for BIP and MOOD, 96.3% (157/163) for DEP and MOOD and 96% (27/28) for ADHD and MOOD. These figures closely resemble the MiXeR estimates for MOOD and SCZ (54%), BIP (54%) and DEP (94%) but are somewhat discrepant from the estimate for MOOD and ADHD (77%). This is likely due to the small number of loci identified in this analysis. Functional annotation analyses for individual analyses are presented in *Supplementary results*.

**Table 1:** Summary of the total number of loci jointly associated with mood instability (MOOD) and each of psychiatric disorders (Psych) schizophrenia (SCZ), bipolar disorder (BIP), major depression (DEP) and attention-deficit hyperactivity disorder (ADHD), number of novel loci in each phenotype, number of loci with concordant effect directions in discovery and replication samples, and number of transdiagnostic loci (overlapping between two or more Psych) identified using the conjunctional false discovery rate (conjFDR) method at a threshold of conjFDR<0.05. The sample sizes of the original Psych GWAS and the number of overlapping loci at genome-wide significance in the original GWAS (5xp<10^−8^) are provided for comparison.

| Psych | Psych GWAS (n) | Joint loci with MOOD at ($p < 5 \times 10^{-8}$) | Joint loci with MOOD at (conjFDR < 0.05) | Novel loci in MOOD (n) | Novel loci in Psych (n) | Replicated loci (sign test) (n) | Transdiagnostic loci (n) |
| --- | --- | --- | --- | --- | --- | --- | --- |
| SCZ | 105,318 | 40 | 102 | 71 | 26 | 74 | 41 |
| BIP | 413,463 | 5 | 60 | 42 | 22 | 39 | 35 |
| DEP | 450,619 | 29 | 163 | 140 | 92 | 121 | 38 |
| ADHD | 53,293 | 2 | 28 | 17 | 12 | N/a | 11 |

### Consistency of genetic effects in independent samples

When comparing the effect directions of lead SNPs in discovery and replication samples, there was significant *en masse* sign concordance for SCZ (74/96 concordant; p=4.72e^-8^), BIP (39/56 concordant; p=0.0023) and DEP (121/154 concordant; p=2.63e^-13^). The discrepancy in the number of lead SNPs was due to missing lead SNPs in replication samples (SCZ=6; BIP=4; DEP=9). We did not have access to sufficiently large independent datasets for MOOD or ADHD.

### Identifying and characterising transdiagnostic loci

A total of 53 loci were associated with MOOD and two or more psychiatric disorders (*Supplementary table 9*). Among these, 38 were associated with MOOD and two disorders, 11 with MOOD and three disorders and four with MOOD and all four disorders. Seven transdiagnostic loci had divergent effect directions on psychiatric disorders, but BIP was always concordant with SCZ (n=27) and DEP was always concordant with ADHD (n=18). The distribution of these loci is summarised in *Figure* 2. Loci overlapping across three or more psychiatric disorders are presented in *Table 2*.

**Table 2:**
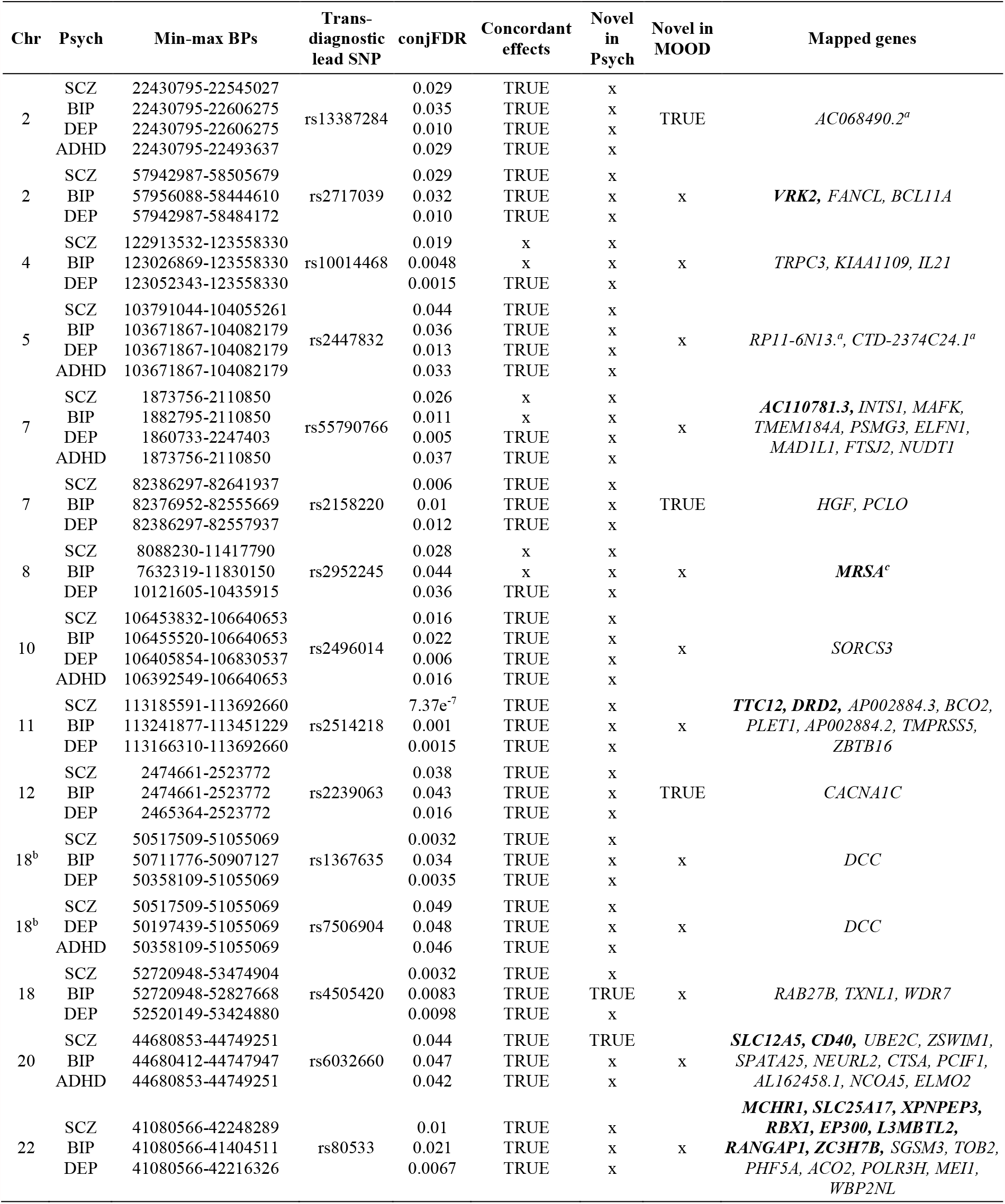
Transdiagnostic loci jointly associated with mood instability (MOOD) and psychiatric disorders (Psych) across three or more disorders. Minimum and maximum base pairs (BPs), “transdiagnostic lead SNPs” and conjunctional false discovery statistics (conjFDR) are presented for each locus. The concordance of effect direction and novelty of a locus for MOOD and each Psych is indicated by “TRUE”. Protein-coding genes mapped to candidate SNPs from each MOOD/Psych analysis are presented. If there were no protein-coding genes mapped to a locus, non-protein coding mapped genes are presented (^a^). Genes mapped by all three mapping strategies (credible genes) are in bold. ^b^Loci are physically overlapping but there was no candidate SNP with conjFDR<0.05 across all four analyses. Locus spans 8p23 inversion region with complex linkage disequilibrium. This biases gene-mapping strategies so only a single mapped gene is presented.

We next identified 1179 genes that were mapped to candidate SNPs from two or more psychiatric disorder/MOOD pairs, 64 of which were mapped by all three strategies (*Supplementary table 7*). *Figure 2* illustrates the chromosomal distribution of the shared loci for each phenotypic pairing alongside mapped genes for each transdiagnostic locus overlapping across three or more disorders. Among these, *VRK2, KIAA1109, AC110781*.*3, PCLO, TMPRSS5* and *EP300* were all mapped to non-synonymous exonic SNPs. Furthermore, *VRK2, AC110781*.*3* and *EP300* were mapped by all three mapping strategies, including eQTLs in the substantia nigra (*VRK2*), caudate, hypothalamus and nucleus accumbens (*AC110781*.*3*) and the cerebellum and hypothalamus (*EP300*). *VRK2* is a serine threonine kinase which has previously been implicated in SCZ, BIP and DEP and plays a role in neuronal proliferation and migration (39). *AC110781*.*3* is a gene of unknown function expressed within 13 different brain tissues, with greatest expression in the cortex, amygdala, and hippocampus (*Supplementary figure 5*). It was also mapped to a locus associated with all four disorders, but with opposite effect directions on SCZ and BIP vs DEP and ADHD. Finally, *EP300* is a histone acetyltransferase implicated in cell proliferation and differentiation. Other notable genes mapped to transdiagnostic loci include the dopamine receptor D2 gene (*DRD2*), the calcium channel voltage-gated channel subunit *CACNA1C*, and the neuron specific potassium/chloride transporter *SLC12A5*.

**Figure 2:**
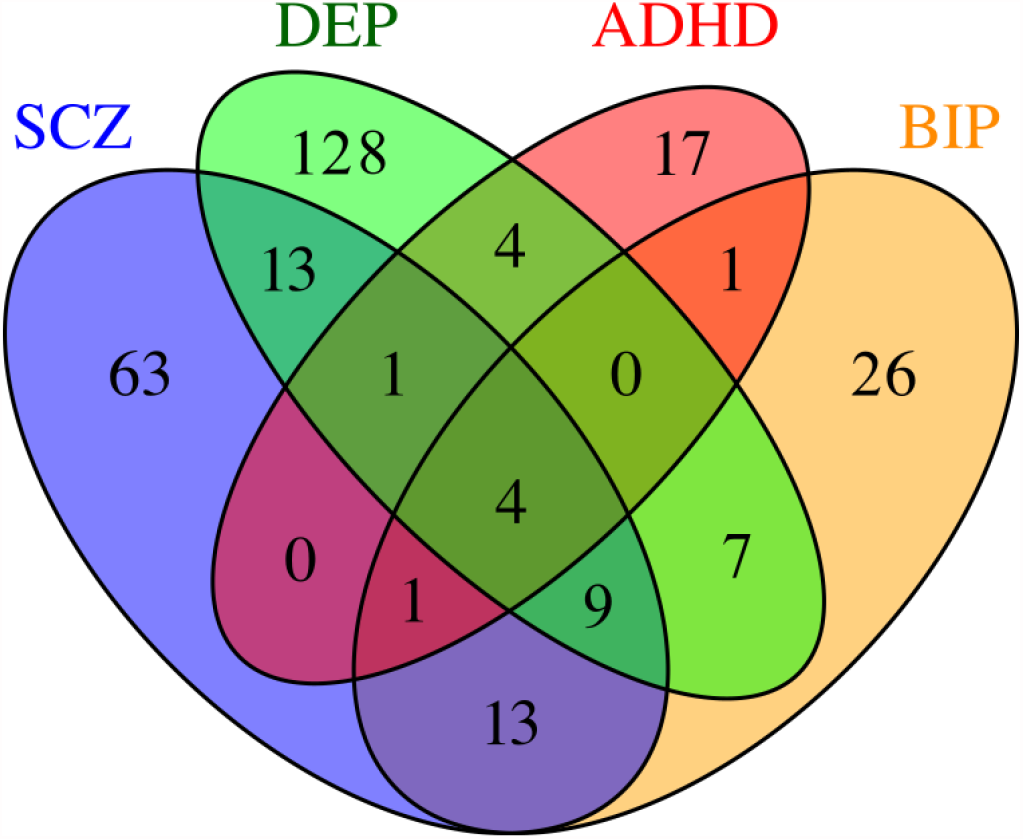
The distribution of transdiagnostic mood instability loci. Venn diagram showing the numbers of diagnosis-specific and transdiagnostic loci across each MOOD and psychiatric disorder conjFDR analysis (SCZ=schizophrenia, BIP= bipolar disorder, DEP=major depression, and ADHD=attention-deficit hyperactivity disorder).

**Figure 3:**
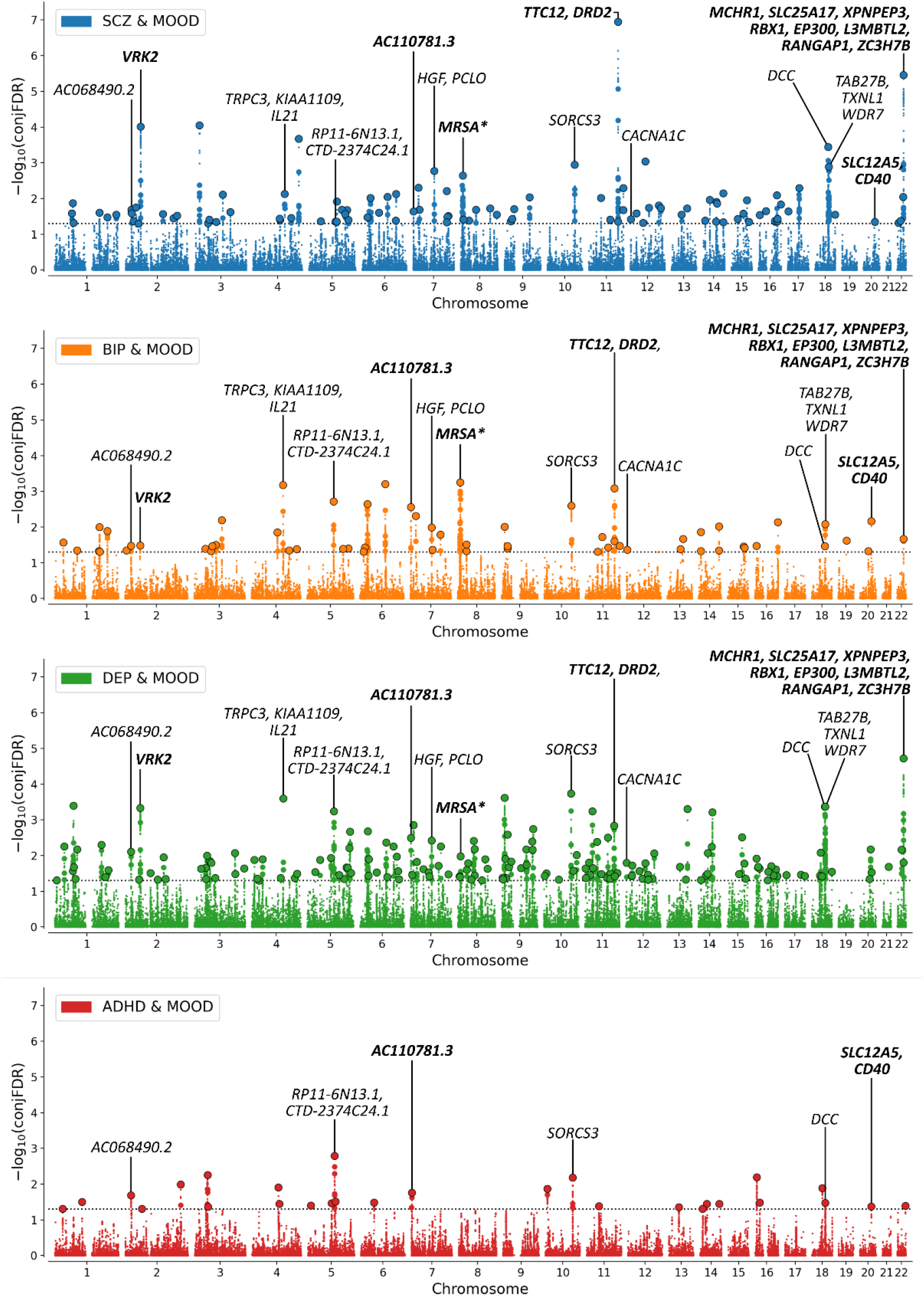
Manhattan plot showing -log10 transformed conjunctional FDR (conjFDR) values for mood instability (MOOD) and ***a)*** schizophrenia (SCZ, blue) ***b)*** bipolar disorder (BIP, orange) ***c)*** major depression (DEP, green) and ***d)*** attention-deficit hyperactivity disorder (ADHD, red) for each SNP (y axis) against chromosomal position (x axis). The dotted line represents conjFDR<0.05 significance threshold. Black circles represent lead SNPs. Lead SNPs from transdiagnostic loci across three or more disorders are annotated with mapped genes. N.B. Not all mapped genes for each locus are presented due to space limitations. Credible genes (bold) were prioritised followed by protein-coding genes and then non-protein-coding genes. * Locus spans 8p23 inversion region with complex linkage disequilibrium. This biases gene-mapping strategies so only a single mapped gene is presented.

A single gene-set, “Synapse organization” was significantly enriched with mapped genes from all four analyses. There were a further 6 gene-sets enriched with mapped genes for MOOD and each of SCZ, BIP and DEP, although there was extensive overlap in genes across the different gene-sets. All gene-sets besides “Neuron part” were either directly or indirectly related to synaptic structure (“Synapse”, “Synapse part”, “Postsynapse”, “Synaptic membrane”, “cell projection part”) (*Supplementary table 8*), Interestingly, when linking mapped genes from each gene-set back to their associated genomic locus, there was a divergent pattern of effect directions with SCZ (51.4-61.8%) and BIP (58.3-70.6%) showing a pattern of mixed effect directions with MOOD, while DEP (97.4-100%) and ADHD (100%) were almost entirely concordant. This is consistent with the patterns of effect directions estimated by MiXeR and observed in jointly associated loci identified by conjFDR (*Supplementary table 10*). This indicates that the divergent pattern of effect directions persists at the level of specific gene-sets.

## DISCUSSION

In this combined GWAS analysis, we used MiXeR to reveal extensive polygenic overlap between MOOD and each of SCZ, BIP, DEP and ADHD despite divergent patterns of genetic correlations. A large proportion of the genetic variants linked to psychiatric disorders also influence MOOD, but the number of shared and trait specific variants and the balance of protective and risk-enhancing variants differ across diagnostic groups. Using conjFDR, MOOD was jointly associated with SCZ at 163 loci, BIP at 60 loci, DEP at 163 loci and ADHD at 28 loci, representing 259 unique genomic loci jointly associated with MOOD and psychiatric disorders. Of these, 220 were novel in MOOD and 152 were novel in psychiatric disorders. Replication analysis provided evidence of consistent genetic effects in independent SCZ, BIP and DEP samples. We identified 53 transdiagnostic loci that were overlapping across MOOD and two or more psychiatric disorders, implicating 1179 putative transdiagnostic genes with an apparent convergence on synaptic gene-sets, although a divergent pattern of effect directions persisted within shared gene-sets. These findings have implications for how the genetic risk of mental-health related traits is conceptualized and suggests differences in the neurobiological basis of MOOD across different psychiatric disorders, including the possibility of genetically influenced sub-groups of patients with more or less prominent MOOD. We also highlight genes that are likely to influence MOOD across several diagnoses, indicating high relevance for future *in vitro* and *in vivo* investigation.

Firstly, 55%-97% of disorder associated variants were predicted to influence MOOD, raising questions about the specificity of the genetic architecture of these complex polygenic psychiatric disorders and related traits. Our findings compliment evidence that a large proportion of genetic variants are not unique for a given mental trait or disorder (26,40), but influence multiple mental phenotypes to different degrees. As such, the distinct SNP-based risk profiles for different mental-health related traits are not merely defined by unique non-overlapping sets of genetic variants, but largely accounted for by a set of pleiotropic non-specific genetic variants showing different strengths of association and effect across these phenotypes (26). Although this hypothesis warrants further interrogation, it suggests that novel approaches are needed to account for the substantial pleiotropy we predict in order to robustly distinguish the genetic risk for different mental traits and disorders.^26^ Furthermore, this places emphasis on identifying disease specific variants that may disproportionately affect the development of a specific phenotype, individually and/or collectively, to inform precision medicine approaches in psychiatry.

Secondly, MOOD has gained interest due to its prevalence across diagnoses and its prominent neurobiological basis (14), implying that it may represent a novel treatment target (2,14). To some extent, this is supported by the large degree of shared genomic loci and corresponding mapped genes identified. However, there were differences in genetic correlations and effect directions of shared loci, with stronger positive correlations and higher proportions of loci with concordant effects in DEP and ADHD compared to weak correlations and lower proportions of loci with concordant effects in SCZ and BIP. This pattern persisted within specific gene-sets identified across multiple analyses. This implies that there may be mechanistic differences in MOOD across the four psychiatric disorders. It is important to note that this measure only reflects one aspect of MOOD, which may explain the lack of correlation with SCZ and BIP, particularly given MOOD’s strong clinical association with BIP. Nonetheless, it is tempting to speculate that MOOD experienced in DEP and ADHD has a similar neurobiological relationship whereas MOOD in BIP and SCZ may reflect a different underlying aetiological mechanism. This is relevant as such differences may limit the potential for transdiagnostic pharmacological interventions. Alternatively, the current findings are also consistent with subgroups characterised by higher or lower MOOD within diagnostic categories, in line with clinical observations (41). Above all, these findings emphasise the importance of exploring the neurobiological and phenomenological differences in MOOD across diagnostic groups.

To characterise MOOD’s neurobiological underpinnings, we used three gene-mapping strategies to identify credible mapped genes for all jointly associated loci. Among these, *AC110781*.*3* was mapped to a non-synonymous exonic SNP jointly associated with MOOD and all four psychiatric disorders. *AC110781*.*3* is a protein-coding gene of unknown function that is expressed in the cortex, amygdala and hippocampus. In addition to previously being implicated in schizophrenia (42) and risk-taking behaviour (43), we also recently linked *AC110781*.*3* to multiple sleep phenotypes and BIP, DEP and SCZ in an analysis of the genetic overlap between sleep-related phenotypes and psychiatric disorders. This suggests *AC110781*.*3* influences multiple diverse phenotypes and may represent a promising candidate for further *in vitro* and *in vivo* investigation. We also identified several well-established psychiatric risk genes, including *VRK2* (39), *CACNA1C* (44), and *DRD2* (45), although an association with mood instability has not previously been described in the literature. The convergence of transdiagnostic mapped genes on synaptic structure builds on *Ward et al*.’s finding in the primary mood instability GWAS that mapped genes were associated with synaptic transmission.

Finally, while mood instability has a prominent genetic component, it remains influenced by environmental factors (12,18). Future work focusing on gene-environmental interplay, particularly in relation to childhood trauma which has been found to correlate with the development of mood instability (46), would be of high interest.

There were limitations to the current study. Firstly, due to available sample sizes and multi-ancestral differences in LD structure we were unable to include multi-ancestral samples. This is an essential area for improvement in psychiatric genetics. Secondly, due to the small sample size of the most recent borderline personality disorder GWAS (n=2 579), we were unable to include it in the current analysis despite its primacy in MOOD (2). This analysis should be repeated as sample sizes increase to include other relevant disorders, identify more transdiagnostic loci, and validate MiXeR’s predictions. Thirdly, the measure of MOOD was based on a single, binary questionnaire item that did not measure affect intensity, regulation of affect or behavioural sequelae, and did not specify the timeframe (1). This may have contributed to the lack of genetic correlations with SCZ and BIP. Nonetheless, a simple measure was necessary to achieve a large enough sample size to achieve a genome-wide signal. Previous GWAS using more complete measures have substantially smaller sample sizes and failed to identify genome-wide significant loci (47). Moreover, the same binary questionnaire item is associated with BIP and DEP, demonstrating its clinical relevance. Future work with more refined measures is required to understand how these findings relate to other dimensions of MOOD. Finally, differences in sample size affect conjFDR’s power to discover shared loci. This precludes cross-analysis comparisons of the number of loci discovered by conjFDR. The disparity between the number of shared loci discovered and the number of shared variants predicted by MiXeR also indicates that we cannot confidently identify loci “unique” to each mental disorder, since it is possible that the lack of association is due to type II error. This will only be addressed once larger proportions of disorder-influencing variants have been discovered.

In conclusion, we have discovered extensive polygenic overlap between MOOD and psychiatric disorders with divergent patterns of genetic correlation and effect directions. These results support the notion that there are common molecular pathways implicated in MOOD across diagnostic categories, but disorder specific effect size distributions indicate key differences in MOOD’s neurobiological underpinnings across diagnoses.

## Data Availability

All PGC data are available at https://www.med.unc.edu/pgc/download-results/, schizophrenia meta-analysis of PGC and CLOZUK datasets at https://walters.psycm.cf.ac.uk/, height at https://portals.broadinstitute.org/collaboration/giant/index.php/GIANT_consortium_data_files, and FinnGen at https://www.finngen.fi/en/access_results. Full GWAS summary statistics for the 23andMe depression dataset will be made available through 23andMe to qualified researchers under an agreement with 23andMe that protects the privacy of the 23andMe participants. Interested investigators should and reference this paper for more information. Summary statistics for mood instability are available upon request by contacting the corresponding author of the original mood instability GWAS.
The code for all analyses can be accessed at https://github.com/precimed.

https://walters.psycm.cf.ac.uk/

https://portals.broadinstitute.org/collaboration/giant/index.php/GIANT_consortium_data_files

https://www.finngen.fi/en/access_results

## ACKNOWLEDGEMENTS

We would like to thank Dr. Yunhan Chu for her thoughtful contributions to the manuscript. We thank the research participants and employees of the Schizophrenia, Depression and Bipolar Disorder Working Groups of the Psychiatric Genomics Consortium, 23andMe, UK Biobank, and FinnGen for making this research possible. We gratefully acknowledge support from the American National Institutes of Health (NS057198, EB00790), the Research Council of Norway (229129, 213837, 223273), the South-East Norway Regional Health Authority (2017-112), KG Jebsen Stiftelsen (SKGJ-MED-008) and the PGC US Norway Collaboration (RCN# 248980). This project has received funding from the European Union’s Horizon 2020 research and innovation programme under grant agreement No 847776. This research has been conducted using data from UK Biobank, a major biomedical database (Project ID number 27412; www.ukbiobank.ac.uk).

