## Supplementary material for "Genome-wide association analysis reveals extensive genetic overlap between mood instability and psychiatric disorders but divergent patterns of genetic effects"

**Supplementary methods: 	2-4**

**Supplementary results:    	5-6**

**Supplementary figures:	7-10**

**References:	11**

### Supplementary methods

#### Samples

We acquired summary statistics from Ward et al.’s 2019 GWAS of MOOD in the European ancestry subset of the UK Biobank (n=363,705)^1^. The UK Biobank is a large-scale population cohort aged from 39 to 69^2^. Individuals with self-reported DEP, BIP, SCZ, “nervous breakdown”, self-harm, suicide attempt or psychotropic medication use were excluded from the primary analysis to limit the confounding effect of psychiatric diagnosis^1^. 157,039 included participants reported mood instability (43.2%).

The CLOZUK sample comprised patients with confirmed diagnoses of treatment-resistant schizophrenia as reported the clozapine blood-monitoring system^3^. The 23andMe DEP cohort cases and controls were defined by self-report in web-based surveys^4^. The remaining PGC cohorts defined cases using international diagnostic criteria (DSM-IV or ICD-10) for a lifetime diagnosis of SCZ, BIP, DEP or ADHD. Diagnoses were made using structured diagnostic instruments administered by trained interviewers, clinician-administered checklists or review of medical record. Controls were randomly selected from the population and screened for the absence of lifetime psychiatric disorders^5–8^.

Within the BIP cohort, 25,060 had BIP type I, 6,781 had BIP type II, 977 had schizoaffective disorder bipolar type, and 9099 had unspecified BIP^9^. The height sample comprised a meta-analysis of the GIANT consortium (n=~237,103) and UK Biobank (n=~456,426)^10^.

The DEP sample included 59,851 cases who met diagnostic criteria for major depressive disorder, a more severe form of depression defined by the number and severity of symptoms experienced, and 75,607 cases who had self-reported a diagnosis or treatment of clinical depression by a medical professional. While differing definitions of depression have been shown to impact SNP heritability and patterns of genetic correlations with related phenotypes^11^, the weighted mean genetic correlation was 0.76 (s.e.=0.03) across all cohorts which is lower but comparable to an equivalent GWAS in SCZ which did not include self-report samples (0.84, s.e. 0.05). This supports the use of the combined DEP sample

All samples were checked for sample overlap with the UK Biobank cohort. For conditional/conjunctional FDR analysis, which is sensitive to sample overlap, UK Biobank subsamples were excluded from the psychiatric disorders when present. Given height was only included in the MiXeR analysis and MiXeR is not sensitive to overlapping datasets, the inclusion of UK Biobank data in height was acceptable.

#### Replication Samples

The SCZ replication sample comprised 22,778 cases and 35,362 of East Asian descent^12^. The use of samples from different ancestral groups for replication may increase the probability of negative replication due to differences in linkage disequilibrium structure across ancestries. In contrast, positive replication across ancestral groups is unlikely to be biased by different LD structures, and is supported by the strong positive genetic correlation between European and East Asian SCZ samples (r_g_=0.98)^12^.

The FinnGen BIP (n cases = 4,501, n controls = 192,220) and DEP (n cases = 17,794, n controls = 156,611) were derived from inpatient, outpatient and cause of death registries using ICD-codes for bipolar affective disorders and depressive episode or recurrent depressive episode respectively. There were no Finish samples in either the primary BIP or DEP samples.

#### MiXeR Analysis

MiXeR (v1.3) constructs causal mixture models from GWAS summary statistics (<https://github.com/precimed/mixer>)^13,14^. First, MiXeR performs a univariate analysis by modelling the additive genetic effect of allele substitution,$\beta_{i}$, for each SNP, $i$, as a point-normal mixture, $\beta_{i}=\left( 1-\pi_{1} \right)N\left( 0,0 \right)+\pi_{1}N(0, \sigma_{\beta}^{2})$, whereby $\pi_{1}$is the proportion of trait-influencing SNPs (i.e. “causal” SNPs/non-null SNPs/SNPs with true genetic effect beyond linkage disequilibrium), referred to as ‘polygenicity’, and $\sigma_{\beta}^{2}$ is the variance of effect sizes of “trait-influencing” SNPs, referred to as ‘discoverability'. Next, MiXeR combines LD data and allele frequencies for 9,997,231 SNPs from the 1000 Genomes Phase3 data for each SNP, $j$^15^. Using this information, the expected probability distribution of the signed test statistic is computed as $z_{j}=\delta_{j}+\epsilon_{j}=N\sum_{i} \sqrt{H_{i}}r_{ij}\beta_{i}+\epsilon_{j}$, in which $N$ represents sample size, $H_{i}$ represents the heterozygosity of SNP *i*, $r_{ij}$ represents allelic correlation between SNP *i* and *j*, and $\epsilon_{j}\sim N(0, \sigma_{0}^{2})$ represents residual variance. Direct maximization of the likelihood function is used to fit the three parameters, $\pi_{1}, \sigma_{\beta}^{2}, \sigma_{0}^{2}$. The number of “trait-influencing” variants is finally computed as $M\pi_{1}$, in which M = the number of SNPs within the LD reference panel.

In order to estimate polygenic overlap between two traits, MiXeR performs a bivariate analysis which models additive genetic effects as a mixture of four components: 1) SNPs not influencing either trait ($\pi_{0})$; SNPs influencing either 2) the first or 3) the second trait ($\pi_{1}$ and $\pi_{2}$, respectively); and 4) SNPs influencing both traits ($\pi_{12}$, shared component). MiXeR models the shared component as a variance-covariance matrix as $\Sigma_{12}=\left[ \sigma_{1}^{2} {\rho_{12}\sigma}_{1}\sigma_{2} {\rho_{12}\sigma}_{1}\sigma_{2} \sigma_{2}^{2} \right]$ where $\rho_{12}$ represents the genetic correlation (correlation of effect sizes) within the shared component, and $\sigma_{1}^{2}$ and $\sigma_{2}^{2}$ represent the discoverability of the two traits as estimated in the univariate analysis. The genetic correlation is then computed as $r_{g}=\frac{\rho_{12}\pi_{12}}{\sqrt{(\pi_{1}+\pi_{12})(\pi_{2}+\pi_{12})}}$.

After estimating the size of the shared and unique components, the dice coefficient (DC) was calculated to represent the overall extent of genetic overlap using the formula DC = $\frac{2\pi_{12}}{{\pi_{1}+\pi_{2}+2\pi}_{12}}$.

All point estimates and standard deviations were computed by conducting 20 iterations with 2 million random SNPs followed by random pruning at an r^2^ threshold of 0.8, (i.e. ~600K SNPs per iteration).

To identify insufficiently powered analyses, the Akaike information criterion ($AIC=2k-2lnL$) was used, in which $k$ represents the number of free parameters, $L$ represents the value of the likelihood function, and $n$ represents the number of SNPs used in the optimization procedure. The difference in $AIC$ between the complete model ($k=3$) and a reduced model ($k=2$). The reduced models are generated by constraining the shared component ($\pi_{12}$) to either the smallest (minimum overlap required to generate observed genetic correlation) or largest possible value (complete genetic overlap) which are calculated as $\pi_{12}^{min}=r_{g}\sqrt{\pi_{1}^{u} \pi_{2}^{u}}$ and $\pi_{12}^{max}=min(\pi_{1}^{u}, \pi_{2}^{u})$, respectively). A positive AIC value indicates that the GWAS summary statistics have enough information to distinguish the complete model’s estimate of polygenic overlap versus the constrained models with minimal ($\pi_{12}^{min}$) and maximum ($\pi_{12}^{max}$) polygenic overlap.

#### Conditional QQ plots

Conditional Q-Q plots compare the enrichment of the association of all SNPs with a primary phenotype (e.g. MOOD) with enrichment for the same phenotype within three subsets of SNPs stratified by the strength of their association to a second phenotype (e.g. SCZ), set at p<0.1, p<0.01 and p<0.001.

The data points on the QQ plot are weighted according to the LD structure around the corresponding SNP. We used n=200 iterations of random pruning with an LD threshold r2=0.1 to define LD-blocks throughout the genome. For each iteration, only one SNP from each block was selected to contribute to the p-value distribution statistics.

#### Conjunctional FDR analysis

Test-statistics for the primary phenotype were re-ranked according to the significance of their association with the secondary phenotype enabling the estimation of the posterior probability that a SNP has no association with the primary trait, given that the p-values for that SNP in both the primary and conditional traits are as small as or smaller than the observed p-value. We reversed primary and secondary phenotypes to calculate the inverse condFDR value.

The conjunctional FDR statistic is then computed as the maximum of the conditional and inverse conditional FDR statistic since this represents a conservative estimate of the FDR for the association with both traits. This thus provides an estimate for the posterior probability that a SNP is null for either trait or both, given that the p-values for both phenotypes are as small as or smaller than the p-values for each trait individually. Please refer to the following original publication and review article for more detail of the conditional/conjunctional FDR method ^16,17^.

Genomic regions with complex LD are liable to confound FDR estimation and so the major extended histocompatibility complex (MHC) and 8p23.1 regions were excluded from this analysis.

#### Genomic loci definition and novelty checking

According to the FUMA protocol, physically overlapping lead SNPs (<250kb apart) were merged. The borders of each genomic loci were delineated by identifying the region comprising all SNPs in linkage disequilibrium (LD) (r^2^≥0.6) with each significant independent SNP.

Novel loci were determined by cross-referencing identified loci with previous GWAS and other relevant studies.^3,5,7,8,12,18–22^

#### Replication of conjFDR loci in independent samples

Due to small effect sizes observed in GWAS, the probability of successfully replicating individual genomic loci at genome-wide significance is low, particularly when the sample sizes for the replication cohorts are small. In line with recent GWAS^23,24^, we therefore tested for *en masse* sign concordance and counted nominally significant lead SNPs in independent samples. First, effect sizes and p-values for all lead SNPs were extracted from the replication cohorts. Six, 4 and 9 lead SNPs were not present in the SCZ, BIP and DEP cohorts respectively, and so were dropped. We next compared the effect directions in the discovery and replication cohorts. After counting the number of lead SNPs with concordant effects in the discovery and replication cohorts, we employed a one-sided exact binomial test of significant against the null hypothesis that 50% of loci have concordant effects.

#### Functional annotation

CADD scores: The combined annotation dependent depletion scores predict how deleterious the SNP effect is on protein structure/function. >12.37 is considered predictive of pathogenicity.

RegulomeDB scores: predict the likelihood that a given SNP possesses a regulatory function. The scores relate to the following features related to the probability of the SNP having a regulatory function (TF = transcription factor):

| 1a | eQTL + TF binding + matched TF motif + matched DNase Footprint + DNase peak |
| --- | --- |
| 1b | eQTL + TF binding + any motif + DNase Footprint + DNase peak |
| 1c | eQTL + TF binding + matched TF motif + DNase peak |
| 1d | eQTL + TF binding + any motif + DNase peak |
| 1e | eQTL + TF binding + matched TF motif |
| 1f | eQTL + TF binding / DNase peak |
| 2a | TF binding + matched TF motif + matched DNase Footprint + DNase peak |
| 2b | TF binding + any motif + DNase Footprint + DNase peak |
| 2c | TF binding + matched TF motif + DNase peak |
| 3a | TF binding + any motif + DNase peak |
| 3b | TF binding + matched TF motif |
| 4 | TF binding + DNase peak |
| 5 | TF binding or DNase peak |
| 6 | other |

Minimum chromatin state scores: the minimum chromatin state across 127 tissues at a given SNP locus. Scores of 1-7 are suggestive of open chromatin state ^25^.

#### Data availability

All PGC data are available at <https://www.med.unc.edu/pgc/download-results/>, SCZ meta-analysis of PGC and CLOZUK datasets at <https://walters.psycm.cf.ac.uk/>, height at <https://portals.broadinstitute.org/collaboration/giant/index.php/GIANT_consortium_data_files>, and FinnGen at <https://www.finngen.fi/en/access_results>. Full GWAS summary statistics for the 23andMe DEP dataset will be made available through 23andMe to qualified researchers under an agreement with 23andMe that protects the privacy of the 23andMe participants. Interested investigators should email [](https://mail.ucsd.edu/owa/redir.aspx?C=KkMl4ROy8xUxV5VcKTOPY0E7jYvTzgjfk2VwJUMjCmkE5-x33hzVCA..&URL=mailto%3adataset-request%4023andme.com) and reference this paper for more information. Summary statistics for MOOD are available upon request by contacting the corresponding author of the original MOOD GWAS.^1^

### Supplementary results

#### Univariate MiXeR analysis

We first applied univariate MiXeR to describe mood instability’s polygenic architecture relative to psychiatric disorders. MiXeR estimated SNP-based heritability was 0.07, slightly lower than the BOLT-LMM estimated heritability reported in the original GWAS (0.09) (*Supplementary table 1*).(1) Approximately 10.4K SNP variants were predicted to explain 90% of mood instability’s SNP heritability compared to 9.7K, 8.1K, 14K, 5.6K and 4K for SCZ, BIP, DEP, ADHD and height respectively. This reveals mood instability’s extensive polygenicity, comparable in scale to the polygenic architectures previously described in psychiatric disorders.^14^ Given its low heritability and high polygenicity, mood instability was also found to have low SNP discoverability similar to that of DEP, with a sample size of over 100 million required for >90% of all mood instability influencing SNPs to meet genome-wide significance. *Supplementary figure 3* illustrates these findings relative to psychiatric disorders and in the context of current GWAS sample size.

#### Bivariate MiXeR analysis

The observed conditional qq plots closely matched the modelled conditional qq plots, indicating good model fit (*Supplementary figure 4*). For MOOD and height, there were AIC increases when comparing the model with both minimum and maximum possible overlap (*Supplementary table 1*). This indicates that the analysis was sufficiently powered to provide an estimate that could be differentiated from both minimum and maximum possible overlap. For each of SCZ, BIP and ADHD with MOOD, the AIC increased when comparing the model with minimum possible overlap but decreased when comparing with maximum possible overlap. This indicates that the analysis was not sufficiently powered for the model to be distinguished from maximum possible overlap. This means that the SCZ/BIP/ADHD and MOOD estimate may be interpreted as a lower limit of overlap but may underestimate the shared component. For MOOD and DEP, AIC increased when comparing the model with maximum possible overlap but decreased when compared to minimum possible overlap. This indicates the model was distinguishable from the maximum possible overlap but not from minimum possible overlap and so may be interpreted as an upper limit of overlap but may overestimate the shared component.

#### Visualising cross-trait enrichment

Conditional QQ plots of mood instability conditional on each psychiatric disorder demonstrated stepwise increments in enrichment of SNPs associated with mood instability as a function of their association with each of SCZ, BIP, DEP and ADHD (*Supplementary figure 1*) and vice versa (*Supplementary figure 2*). These findings are consistent with the extensive polygenic overlap predicted by MiXeR and supported the use of conjFDR to identify the individual shared loci.

#### ConjFDR and functional annotation

We mapped candidate SNPs to putative causal genes by 3 independent gene-mapping strategies. A total of 947, 676, 1587 and 210 genes were mapped across all four analyses for MOOD and SCZ, BIP, DEP and ADHD respectively, 145 of which were mapped by all three gene mapping strategies (*credible mapped genes*) (*Supplementary table 7*). Mapped genes were overrepresented in 121 gene-sets for MOOD and SCZ, 33 for MOOD and BIP, 59 for MOOD and DEP and 6 for MOOD and ADHD (*Supplementary table 8*).

SCZ and MOOD were jointly associated with 102 loci at conjFDR<0.05, 25 of which possessed conjFDR<0.01. Four lead SNPs possessed CADD scores > 12.37, indicating a high probability of pathogenicity (rs111236099, rs56403421, rs3735440, rs61937595) (*Supplementary table 2*). We next mapped 1370 genes to all candidate SNPs (*Supplementary table 3)*, 442 of which were mapped by position, 443 by eQTL resources, 904 by chromatin interaction, and 76 were mapped by all three strategies, representing a subset of credible genes *(bold)* (*Supplementary table 7*). Gene-set analysis of all mapped genes identified 26 cellular components, 94 biological processes and one molecular function (*Supplementary table 8*).

BIP and MOOD were jointly associated with 60 loci at conjFDR<0.05, 15 of which possessed conjFDR<0.01. Four lead SNPs had CADD scores >12.37 (rs1746662, rs12188417, rs2567377, rs2717039) (*Supplementary table 2*). We mapped 1014 genes to all candidate SNPs (*Supplementary table 4)*, 324 of which by position, 280 by eQTL and 710 by chromatin interaction, and 54 genes mapped by all three strategies *(bold)* (*Supplementary table 7*). Gene-set analysis of all mapped genes identified 24 cellular components, eight biological processes and one molecular function (*Supplementary table 8*).

163 loci were jointly associated with DEP and MOOD at conjFDR<0.05, 45 of which possessed conjFDR<0.01. Fourteen lead SNPs possessed CADD scores >12.37 (*Supplementary table* 2). We mapped 1814 genes to all candidate SNPs associated with DEP and MOOD (*Supplementary table 5*). 797 were mapped by position, 435 by eQTL resources, 1637 by chromatin interaction, and 144 were mapped by all three strategies *(bold)* (*Supplementary table 7*). Gene-set analysis of all mapped genes identified 13 cellular components, 37 biological processes and nine molecular function (*Supplementary table 8*).

Twenty-eight loci were jointly associated with ADHD at conjFDR<0.05, four of which possessed conjFDR<0.01. Two lead SNPs had CADD scores >12.37 (rs2567377, rs28535523) (*Supplementary table* 2). We mapped 294 genes to jointly associated candidate SNPs (*Supplementary table 6*). 119 were mapped by position, 53 by eQTL, 234 by chromatin interaction, and 22 were mapped by all three strategies *(bold)* (*Supplementary table 7*). Gene-set analysis of all mapped genes identified three biological processes and two molecular function (*Supplementary table 8*).

### Supplementary figures

**Supplementary figure 1:** Power plot illustrating percentage of variance explained by genome-wide significant SNPs (y-axis) at a given sample size (x-axis) for each of MOOD, SCZ, BIP, DEP, ADHD and height. The sample size required to discover 50% variance for each phenotype is represented by the dot. The current sample size for each phenotype is represented by a triangle, and the estimated variance (%) based on current sample size is listed in parentheses in the legend.

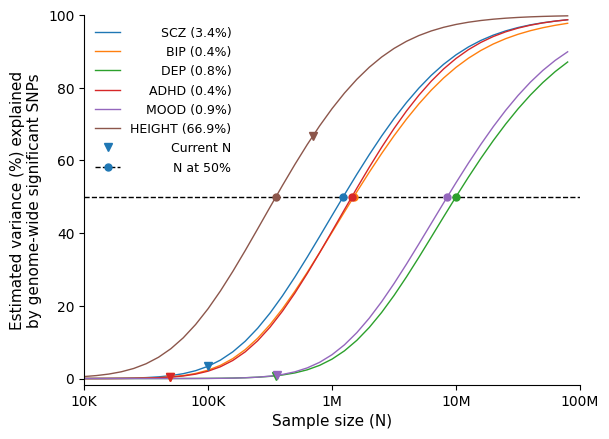

**Supplementary figure 2**: Supplemental MiXeR figures for mood instability (MOOD) and schizophrenia (SCZ), bipolar disorder (BIP), major depression (DEP), attention-deficit hyperactivity disorder (ADHD) and height. On the left, conditional QQ plots of observed versus expected -log_10_ p-values in the primary trait as a function of the significance of the association with the secondary trait at the level of all SNPs ( blue lines), p ≤ 0.1 (orange lines), p ≤ 0.01 (green lines) and p ≤ 0.001 (red lines). Dotted lines indicate model predictions for each stratum. Black dotted line is the expected Q-Q plot under the null hypothesis (no SNPs associated with the phenotype). Points on the Q-Q plot are weighted according to LD structure, using n=64 iterations of random pruning at an LD threshold r2 = 0.1. On the right, likelihood of the MiXeR Model. Negative log-likelihood of the bivariate fit as a function of 𝜋_12_M (i.e. number of shared causal variants). The remaining parameters of the model were constrained to their fitted values.

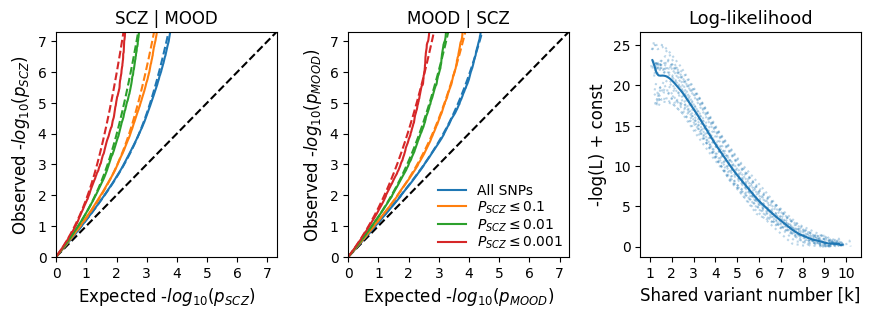

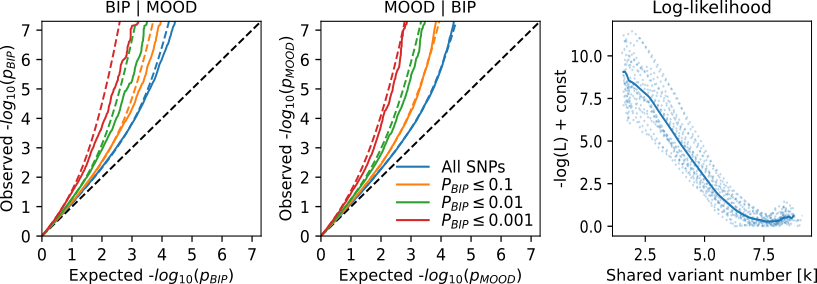

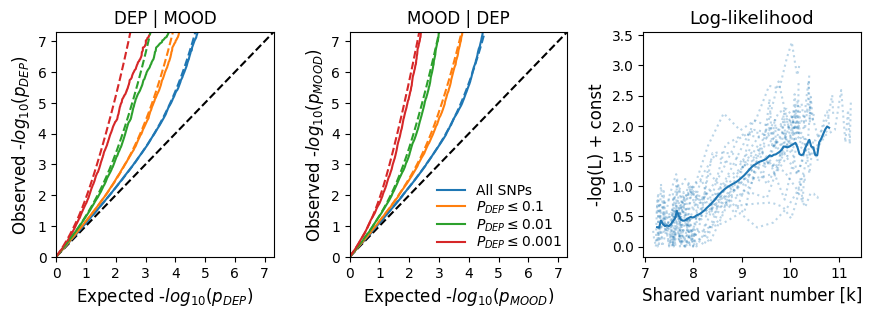

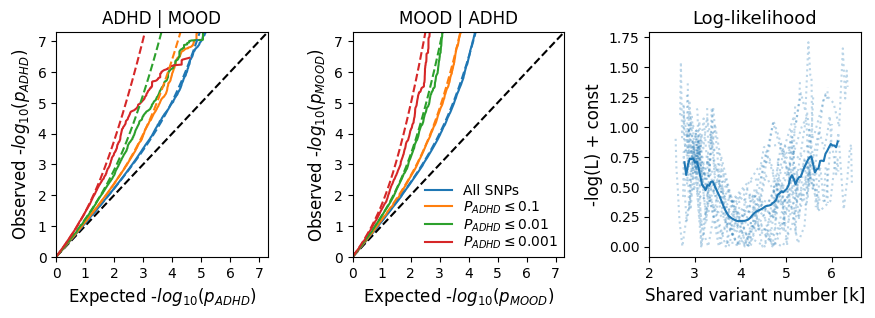

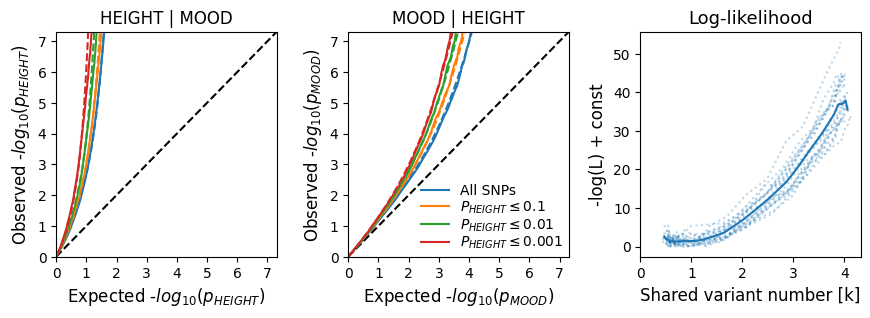

**Supplementary figure 3**: Cross-trait enrichment of SNPs associated with mood instability (MOOD) dependent on their association with schizophrenia (SCZ)**,** bipolar disorder (BIP), major depression (DEP), and attention-deficit hyperactivity disorder (ADHD). Conditional Q-Q plots of nominal versus empirical -log_10_p values for MOOD as a function of the significance of their association with **a)** SCZ, **b)** BIP, **c)** DEP, and **d)** ADHD at the level of all SNPs, p<0.1, p<0.01 and p<0.001 respectively.

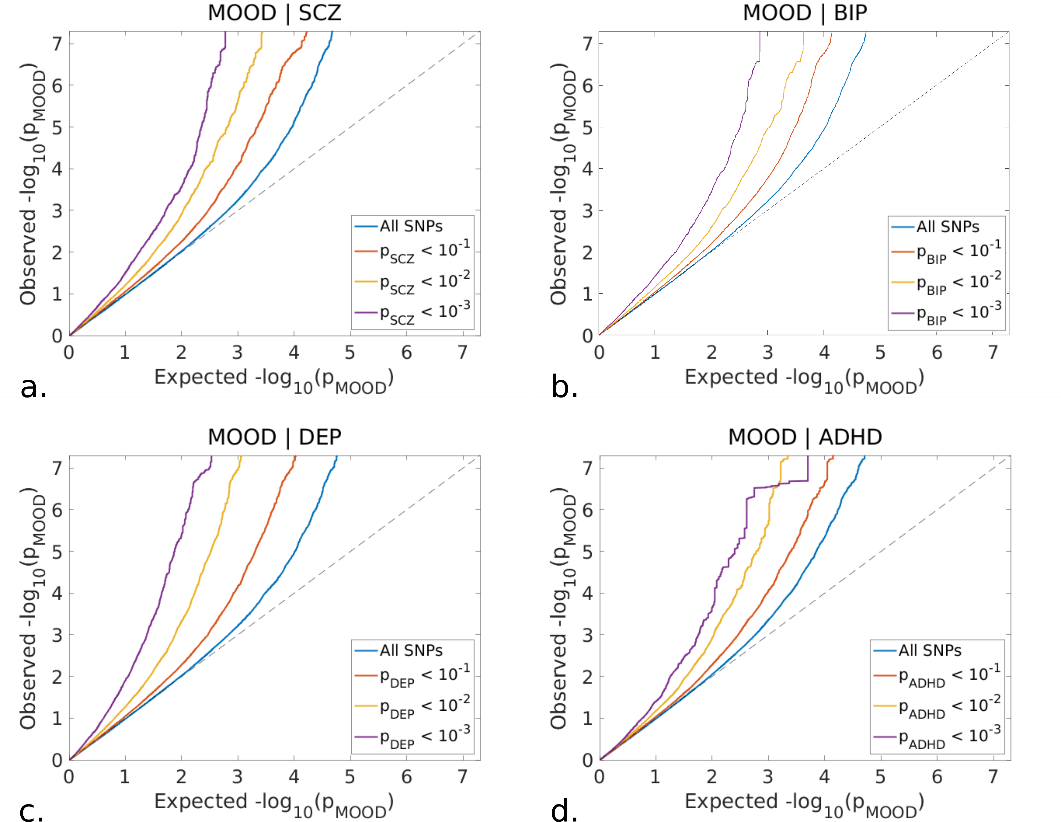

**Supplementary figure 4:** Cross-trait enrichment of SNPs associated with schizophrenia (SCZ), bipolar disorder **(**BIP)**,** major depression **(**DEP) and attention-deficit hyperactivity disorder (ADHD) dependent on their association with mood instability (MOOD). Conditional Q-Q plots of nominal versus empirical -log_10_p values for **a)** SCZ, **b)** BIP, **c)** DEP and **d)** ADHD as a function of the significance of their association with MOOD at the level of all SNPs, p<0.1, p<0.01 and p<0.001 respectively.

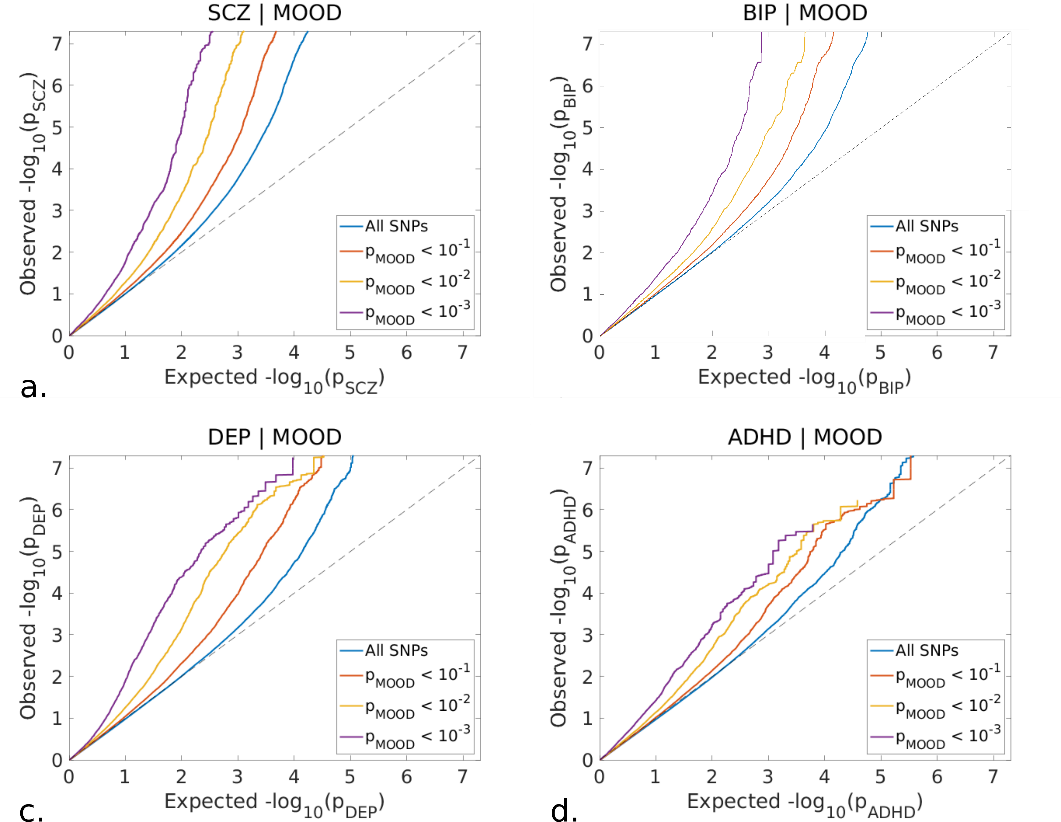

**Supplementary figure 5**: Gene expression for AC110781.3 (ENSG00000176349.11) across selected brain tissues using GTEx V8 [https://gtexportal.org]. Expression values are shown as log_10_TPM (Transcripts Per Million) calculated from a gene model with isoforms collapsed to a single gene. Box plots are shown as median and *25*th and *75*th percentiles. Points are displayed as outliers if they are above or below 1.5 times the interquartile range. Tissues are sorted according to the median value.

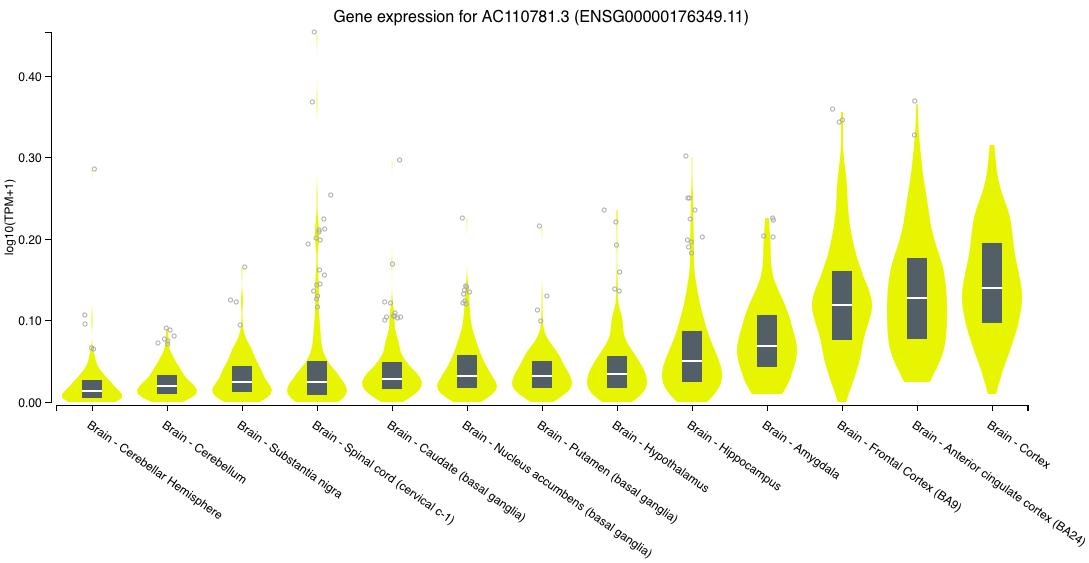
